# Profiles of Distress in OCD: A Cluster Analysis Exploring Associations with Global and Facet-Specific Anhedonia

**DOI:** 10.64898/2025.12.01.25341370

**Authors:** Brian A. Zaboski, Katherine Jones, Elizabeth F. Mattera, Christopher Pittenger, Helen Pushkarskaya

**Author notes:** This work is funded by the State of Connecticut through its support of the Ribicoff Research Facilities at the Connecticut Mental Health Center. This work reflects the views of the authors and not those of the State of Connecticut. In the past three years, BAZ has consulted with Biohaven Pharmaceuticals and received royalties from Oxford University Press; CP has consulted for Biohaven Pharmaceuticals, Freedom Biosciences, Transcend Therapeutics, UCB BioPharma, Mind Therapeutics, Ceruvia Biosciences, F-Prime Capital Partners, and Madison Avenue Partners; has received research support from Biohaven Pharmaceuticals, Freedom Biosciences, and Transcend Therapeutics; owns equity in Alco Therapeutics, Mind Therapeutics, and Lucid/Care; receives royalties from Oxford University Press and UpToDate; and holds pending patents on pathogenic antibodies in pediatric OCD and on novel mechanisms of psychedelic drugs. None of these relationships are related to the current paper.

## Abstract

**Background:** Obsessive-compulsive disorder (OCD) is clinically heterogeneous. Its relationship with anhedonia—a diminished capacity for pleasure—remains unclear, with conflicting findings in the literature. This study identified symptom profiles in OCD to characterize their association with anhedonia.

**Methods:** A k-means cluster analysis was performed on 227 adults with OCD using ten clinical variables, excluding direct measures of anhedonia. The resulting clusters were validated against unidimensional severity measures to confirm their unique construct validity and were then compared on external anhedonia measures.

**Results:** Three distinct and robust profiles emerged: "High Global Distress and OCD" (HGD-OCD; *n* = 62), "Intermediate Overall Severity" (IOS; *n* = 100), and a "Low Symptom Profile" (LSP; *n* = 65). Validation analyses confirmed these profiles represent a multidimensional construct of distress not reducible to depression or OCD severity alone. Anhedonia was strongly associated with these profiles; its severity was highest in the HGD-OCD cluster, intermediate in the IOS cluster, and virtually absent in the LSP cluster. Item-level analyses revealed unique anhedonia signatures, with the IOS profile showing a complex pattern of both impaired and preserved hedonic domains.

**Conclusion:** A data-driven approach effectively stratifies the heterogeneity of distress in OCD. These holistic profiles offer a clinically meaningful alternative to unidimensional measures by revealing distinct, facet-specific manifestations of anhedonia. This framework provides a crucial step toward developing more targeted, personalized interventions for anhedonia in OCD.

---

Obsessive-compulsive disorder (OCD) is characterized by intrusive, unwanted thoughts (obsessions) and repetitive, ritualistic behaviors or mental acts (compulsions) aimed at reducing distress or preventing a feared outcome (American Psychiatric Association, 2022). Affecting 1-3% of the population (Kessler et al., 2012; Ruscio et al., 2010), OCD markedly impairs social, occupational, and personal functioning, underscoring the need for a deeper understanding of its multifaceted nature to refine etiological models and improve treatment efficacy. OCD is characterized by substantial heterogeneity, spanning multiple symptom domains, ages of onset, psychiatric and general health comorbid conditions, physical symptoms, and symptom severity (Mataix-Cols et al., 2009).

Anhedonia—the diminished capacity to experience pleasure (American Psychiatric Association, 2022)—has garnered increasing attention as a transdiagnostic symptom relevant to numerous psychiatric conditions, including OCD (Moore, 2021; Pizzagalli, 2022). Anhedonia is a core feature of major depressive disorder (MDD) and is prominent in psychotic disorders (Barch et al., 2017). Its role in OCD is an area of growing investigation. For example, studies report clinically significant anhedonia in approximately 28% of individuals with OCD (*N* = 113; Abramovitch et al., 2014), with other investigations noting rates as high as 42% (*N* = 77; Moore, 2021). A larger investigation (*N* = 227) found 14.5% meeting criteria for clinically significant anhedonia (Zaboski et al., 2025). This wide variability suggests that anhedonia is a relevant feature for a subset of patients, but its relationship with OCD is complex and not uniform.

Interpreting these findings is complicated by the fact that OCD rarely occurs in isolation. It is frequently accompanied by high rates of comorbid depression and anxiety, which together contribute to a broader syndrome of general distress (Fontenelle et al., 2010; Mataix-Cols et al., 2005). Moreover, similar OCD symptom presentations may relate differently to distress levels because of different resilience capacities (Belhan Çelik et al., 2025). This pattern aligns with contemporary models of psychopathology which posit a general, transdiagnostic factor—the *’p-factor’*—that represents an individual’s overall susceptibility to mental illness (Caspi et al., 2014). We propose that the conflicting findings in the anhedonia literature—specifically, the debate over whether it is an independent feature of OCD or an artifact of comorbid depression (Abramovitch et al., 2014; Pushkarskaya et al., 2019; Zaboski et al., 2025)—are a direct result of studies failing to account for this broader syndrome of general distress. Variable-centered approaches, which treat OCD as a monolithic group, obscure potential interactive or synergistic effects of co-occurring symptoms and mask the underlying clinical reality that different patients experience fundamentally different patterns of overall distress (Lochner & Stein, 2003, 2006).

To overcome this limitation, this study builds on recent computational efforts (Zaboski et al., 2024) with a data-driven approach to identify distinct distress profiles in individuals with OCD. We had two primary objectives: First, we used a k-means approach to identify multidimensional symptom profiles in adults with OCD and to evaluate their construct validity, hypothesizing that these profiles would capture a higher-order construct of distress not reducible to more domain-specific measures of depression, anxiety, or OCD severity. Second, we hypothesized that anhedonia severity would systematically vary across these profiles and that this association would remain significant after controlling for depression and OCD severity. Finally, by examining facet-level patterns of anhedonia, we aimed to clarify *how* it manifests and *for whom* it constitutes a clinically meaningful symptom domain.

## Methods

### Participants

The Yale University Institutional Review Board granted ethical approval for this work. Participant recruitment was conducted through national and local outreach via the Yale OCD Research Clinic. The study population comprised adult individuals (18 – 70 years) diagnosed with primary or co-primary Obsessive-Compulsive Disorder (OCD). Diagnostic confirmation of OCD involved a multi-step process: (1) structured assessment using either the Mini-International Neuropsychiatric Interview (Sheehan et al., 1998) or the Diagnostic Interview for Anxiety, Mood, and OCD and Related Neuropsychiatric Disorders (Tolin et al., 2018); (2) diagnostic validation by a study-affiliated, board-certified psychologist or psychiatrist; and (3) a score of ≥ 16 on the Yale Brown Obsessive-Compulsive Scale (Y-BOCS; Goodman et al., 1989).

These subjects were drawn from the general recruitment flow of the Yale OCD Research Clinic. Standard inclusion and exclusion criteria for our Clinic were applied. These included any significant medical condition with known central nervous system effects (e.g., seizure disorders, cerebrovascular events, poorly controlled thyroid disease, unmanaged hypertension), medical instability, or a history of traumatic brain injury leading to a loss of consciousness for over 30 minutes. Further psychiatric contraindications for participation were the presence of another primary diagnosis of greater clinical significance than the OCD, active suicidal ideation with a plan/intent, past psychosurgery, or current mania or psychosis. Participants with active substance use disorders within the last 6 months were excluded, with exceptions made for mild cannabis, alcohol, or tobacco use disorders.

Upon completion of the informed consent procedure, participants engaged in self-report data submission using REDCap (Harris et al., 2009, 2019), a secure, web-based data capture system adhering to HIPAA regulations. All individuals received $40 for the completion of the questionnaires.

## Measures

### Snaith-Hamilton Pleasure Scale

Anhedonia was quantified using the Snaith-Hamilton Pleasure Scale (SHAPS), a 14-item self-report measure (Snaith et al., 1995). Participants reflected on their hedonic capacity in the last few days when responding to statements via a 4-point Likert scale (“Strongly Agree” to “Strongly Disagree”). Representative statements include, “I would enjoy being with my family or close friends,” and “I would be able to enjoy a landscape or a beautiful view.” A dichotomous scoring protocol is employed for the SHAPS, whereby items marked as “disagree” are scored as 1, and those marked as “agree” are scored as 0, yielding a score range of 0 to 14. Scores of 3 or greater represent clinically meaningful anhedonia (Snaith et al., 1995). Psychometric evaluations have confirmed the SHAPS’s high internal consistency (ɑ = 0.82; Nakonezny et al., 2010) and stability over time (test-retest *r* = .70; Franken et al., 2007).

### Beck Depression Inventory-II

Depressive symptoms were assessed using the Beck Depression Inventory-II (BDI-II; Beck et al., 1996), a 21-item self-report questionnaire. Participants use a 4-point Likert scale to rate four statements for each item based on the past two weeks. Total scores can range from 0 to 63. Standardized cutoffs categorize severity as minimal/no depression (0 – 13), mild depression (14 – 19), moderate depression (20 – 28), and severe depression (29 – 63) (Beck et al., 1996). Validation in various populations and clinical settings supports the BDI-II, which shows high internal consistency (ɑ = 0.91; Dozois et al. 1998) and good test-retest reliability (*r* = 0.73 – 0.96; Wang & Gorenstein, 2013).

Following the framework established by Joiner et al. (2003), the General Depression subscale (gBDI) was created with 18 items and the Anhedonic subscale (aBDI) was derived from the remaining items assessing loss of pleasure (item #4), loss of interest (item #12), and loss of interest in sex (item #21). In the present study, the gBDI subscale was used for clustering; the aBDI was excluded from clustering and used for external validation. This ensured that clusters were not driven by variation in anhedonia. Previous studies have substantiated this two-factor conceptualization (Cogan et al., 2024).

### Quality of Life Enjoyment and Satisfaction Questionnaire Short Form

The Quality of Life Enjoyment and Satisfaction Questionnaire Short Form (Q-LES-Q-SF) was administered to measure subjective well-being. This 16-item instrument is a condensed version of the comprehensive 93-item self-report Q-LES-Q (Endicott et al., 1993). Participants reflect on several life domains, including their mood, social interactions, economic situation, and health status over the past week. Each item is rated on a 5-point Likert scale (“very poor” to “very good”). Total scores range from 14 to 70. Higher values correspond to a greater degree of life satisfaction and enjoyment. The Q-LES-Q-SF has excellent internal consistency (ɑ = 0.90) and test-retest reliability (*r* = 0.93; Stevanovic, 2011).

### Dimensional Obsessive-Compulsive Scale

Obsessive-compulsive symptom severity was assessed using the Dimensional Obsessive-Compulsive Scale (DOCS; Abramowitz et al., 2010). This 20-item self-report questionnaire assesses the severity of OCD symptoms across four core dimensions: responsibility for harm, contamination fears, the presence of unacceptable thoughts, and preoccupation with symmetry and completeness. Participants respond to each item using a 5-point Likert scale. Higher scores reflect more severity or impairment for each symptom. Total scores range from 0 to 80, with cut scores exceeding 21 best distinguishing individuals with OCD from non-clinical populations (Abramowitz et al., 2010). The DOCS has high internal consistency (ɑ = 0.90) and moderate test-retest reliability (*r* = 0.66) (Abramowitz et al., 2010). Convergent validity is established by correlations with other measures of OCD severity, including the YBOCS (*r* = 0.54) and the Obsessive-Compulsive Inventory (*r* = 0.69) (Foa et al., 2002; Rapp et al., 2016).

### State-Trait Anxiety Inventory

The State-Trait Anxiety Inventory (STAI) is a 40-item self-report questionnaire to characterize anxiety (Spielberger et al., 1983). Using a 4-point Likert scale, participants report how they feel at the present moment then how they feel in general. Sample items include “I feel at ease” and “I worry too much over something that really doesn’t matter” (Spielberger et al., 1983). Scores range from 20 to 80 for each subscale, with cutoffs greater than 40 (State) and greater than 44 (Trait) indicating clinically significant levels of anxiety (Linde et al., 2022). The STAI demonstrates high internal consistency (ɑ = 0.90; state, ɑ = 0.89; trait) and test-retest reliability. Test-retest correlations are higher for the trait subscale than the state subscale (*r* = 0.88 vs *r* = 0.70; Barnes et al., 2002). Meta-analyses demonstrate convergent validity with several measures of anxiety symptom severity (*r* = 0.59 – .61; Knowles & Olatunji, 2020).

### Obsessive-Compulsive Trait Core Dimensions Questionnaire

The Obsessive-Compulsive Trait Core Dimensions Questionnaire (OCTCDQ) is a 20-item self-report measure used to assesses OCD severity as it pertains to two core motivational dimensions: harm avoidance (HA) and incompleteness (INC) (Summerfeldt et al., 2001, 2014). Sample items include “It takes a long time for me to feel certain about things” and “There are things that I am afraid might happen if I don’t take certain steps to prevent them.” Items are rated on a 5-point Likert scale from 0 (Never applies to me) to 4 (Always applies to me). Total scores range from 0 to 40 for each subscale.

Longer versions of this form have been developed and demonstrate high internal consistency in both non-clinical (ɑ = .89 for HA, ɑ = .88 for INC) and clinical samples (ɑ = .92 for HA, ɑ = .91 for INC; Summerfeldt et al., 2014), as well as good test-retest reliability (*r* = .89, p < .0001; Puccinelli, 2024). Moreover, evidence demonstrates good convergent validity between OCTCDQ scores and OCD severity (Lundström et al., 2024). This lends support to the underlying constructs assessed by the shorter form, which has not itself been extensively validated.

### Physical Symptom Checklist

Physical symptoms were assessed with the Rotterdam Symptom Checklist (RSCL) Physical Symptom Distress subscale (de Haes et al., 2012). This 23-item self-report instrument assesses distress caused by common physical complaints. Participants rate how troubled they have been by an indicated symptom (e.g., lack of appetite, shortness of breath, and nausea) over the past week, on a 4-point Likert scale ranging from 1 (not at all) to 4 (very much). Scores range from 23 to 92, with higher scores indicating more distress. The RSCL is a widely-used measure of physical discomfort, demonstrating adequate reliability (ɑ = 0.60 – 0.83) across physical symptom subscales (Tchen et al., 2002) and convergent validity with measures of pain and discomfort (*r* = 0.5 for both; Pelayo-Alvarez et al., 2013).

### PROMIS Anxiety

The Patient Reported Outcomes Measurement Information System Anxiety (PROMIS-A) Bank v1.0 was used to assess anxiety. The PROMIS-A contains a bank of 55 items, asking respondents to rate them on a 5-point Likert scale reflecting frequency over the past seven days, from 1 (Never) to 5 (Always) (HealthMeasures, 2019). Sample items include, “In the past 7 days, it scared me when I felt nervous,” and “In the past 7 days, I avoided public places or activities.” The measure is administered as a computer adaptive test (CAT), in which the digital system automatically generates additional questions based on previous responses. Participants respond to a minimum of 4 items, and continue until either the standard error between items drops below a threshold of 3.0, or they have answered 12 items (Pilkonis et al., 2011). PROMIS instruments are standardized with T-Scores (*M* = 50). Higher scores reflect higher anxiety; the recommended cutoff indicating mild anxiety is 55 (HealthMeasures, 2019). The PROMIS-A demonstrates good convergent validity with the Hospital Anxiety and Depression Scale (*r* = 0.84; Zigmond & Snaith, 1983) and Generalized Anxiety Disorder-7 (*r* = 0.79; Spitzer et al., 2006; Clover et al., 2022), as well as good test-retest reliability (ICC = 0.78; Clover et al., 2022; van der Willik et al., 2023).

### PROMIS Depression

The Patient Reported Outcomes Measurement Information System Depression (PROMIS-D) Bank v1.0 was used to assess depression in the sample. Like the PROMIS-A, it is administered adaptively, with a bank of 54 items rated on the same Likert scale. Sample items include, “In the past 7 days, I felt that I had nothing to look forward to” and “In the past 7 days, I felt that I was to blame for things.” Participant continuation rules for the PROMIS-D are the same as the PROMIS-A, and T-scores are also used (Pilkonis et al., 2011). The PROMIS Depression item bank demonstrates good convergent validity with established measures of depression such as the Center for Epidemiological Studies Depression Scale (Radloff, 1977) and the Patient Health Questionnaire (Kroenke et al., 2001) (*r* = 0.72 – 0.84; Pilkonis et al., 2014), as well as good test-retest reliability (ICC = 0.81; van der Willik et al., 2023).

### Measure Rationale

Clustering variables were strategically selected to create comprehensive, data-driven profiles of distress among individuals with OCD. This was achieved by including a broad set of 10 clinical measures that captured disorder-specific symptoms (DOCS, OCTCDQ), general psychopathology including depression and anxiety (gBDI, STAI), physical symptom burden (PSC), and overall functioning (QLES-Q-SF). Crucially, direct measures of anhedonia were intentionally excluded from the clustering algorithm to avoid circularity and ensure that any subsequent associations found between the distress profiles and anhedonia were not an artifact of the clustering process itself.

### Analytic Plan

#### Preprocessing

Preprocessing included scoring and data-cleaning procedures applied across all self-report measures. For the Snaith-Hamilton Pleasure Scale (SHAPS), consistent with Snaith et al. (1995), individual items were dichotomized for the calculation of the total score: Responses indicating disagreement with experiencing pleasure (raw scores of 1 or 2 on the original 1– 4 Likert scale) were coded as “1” (an anhedonic response), while responses indicating agreement (raw scores of 3 or 4) were coded as “0” (a hedonic response). These dichotomized item scores were summed to yield the SHAPS total score (range 0 – 14), with higher scores reflecting greater anhedonia. For some descriptive analyses, the original 4-point Likert scale 1 (Strongly Disagree) to 4 (Strongly Agree) for individual SHAPS items was also retained. Based on established cutoffs for the SHAPS total score, participants with a total score of 3 or greater were categorized into an "anhedonia group" for selected descriptive comparisons. Total and/or relevant subscale scores were computed for all other clinical instruments.

For clustering analyses and subsequent characterization, missing data within numerical clinical variables were addressed using K-Nearest Neighbors imputation (with *k* = 5 neighbors) (Emmanuel et al., 2021). Categorical demographic variables (sex, marital status, ethnicity, average yearly household income) were imputed using their respective modes if missing values were present. Missing data ranged from approximately 1 – 10% (see Table 1 for more details).

**Table 1:**
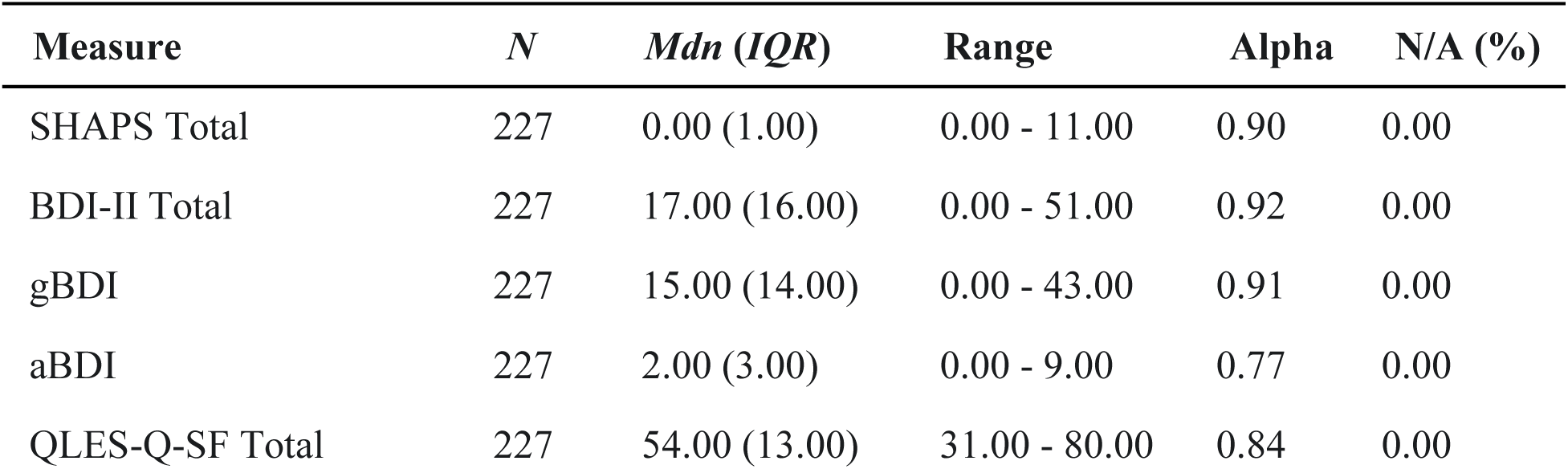
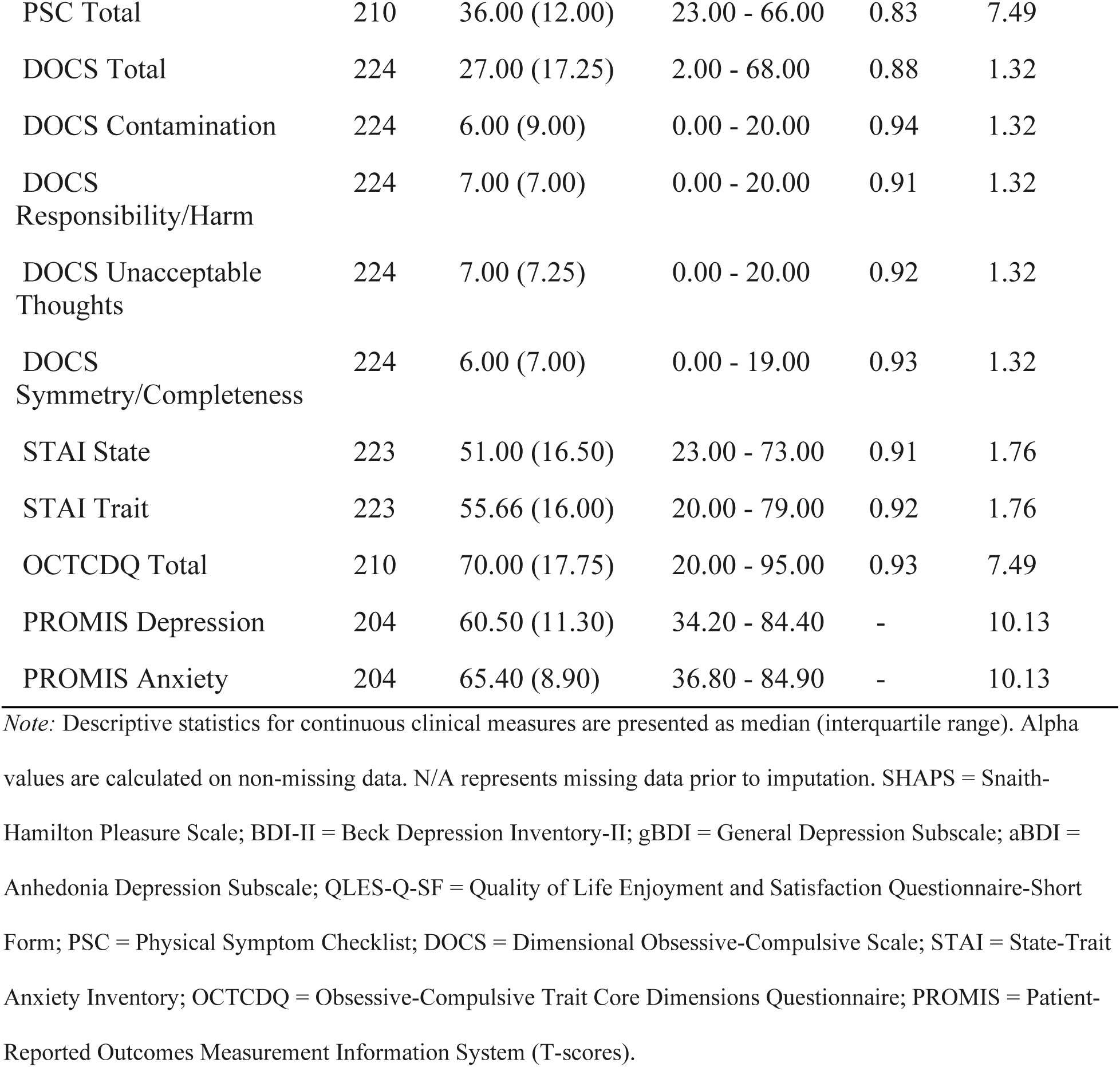
Descriptive Statistics and Internal Consistency of Clinical Measures.

| Measure | *N* | *Mdn (IQR)* | Range | Alpha | N/A (%) |
| --- | --- | --- | --- | --- | --- |
| SHAPS Total | 227 | 0.00 (1.00) | 0.00 - 11.00 | 0.90 | 0.00 |
| BDI-II Total | 227 | 17.00 (16.00) | 0.00 - 51.00 | 0.92 | 0.00 |
| gBDI | 227 | 15.00 (14.00) | 0.00 - 43.00 | 0.91 | 0.00 |
| aBDI | 227 | 2.00 (3.00) | 0.00 - 9.00 | 0.77 | 0.00 |
| QLES-Q-SF Total | 227 | 54.00 (13.00) | 31.00 - 80.00 | 0.84 | 0.00 |
| PSC Total | 210 | 36.00 (12.00) | 23.00 - 66.00 | 0.83 | 7.49 |
| DOCS Total | 224 | 27.00 (17.25) | 2.00 - 68.00 | 0.88 | 1.32 |
| DOCS Contamination | 224 | 6.00 (9.00) | 0.00 - 20.00 | 0.94 | 1.32 |
| DOCS Responsibility/Harm | 224 | 7.00 (7.00) | 0.00 - 20.00 | 0.91 | 1.32 |
| DOCS Unacceptable Thoughts | 224 | 7.00 (7.25) | 0.00 - 20.00 | 0.92 | 1.32 |
| DOCS Symmetry/Completeness | 224 | 6.00 (7.00) | 0.00 - 19.00 | 0.93 | 1.32 |
| STAI State | 223 | 51.00 (16.50) | 23.00 - 73.00 | 0.91 | 1.76 |
| STAI Trait | 223 | 55.66 (16.00) | 20.00 - 79.00 | 0.92 | 1.76 |
| OCTCDQ Total | 210 | 70.00 (17.75) | 20.00 - 95.00 | 0.93 | 7.49 |
| PROMIS Depression | 204 | 60.50 (11.30) | 34.20 - 84.40 | - | 10.13 |
| PROMIS Anxiety | 204 | 65.40 (8.90) | 36.80 - 84.90 | - | 10.13 |

*Note:* Descriptive statistics for continuous clinical measures are presented as median (interquartile range). Alpha values are calculated on non-missing data. N/A represents missing data prior to imputation. SHAPS = Snaith-Hamilton Pleasure Scale; BDI-II = Beck Depression Inventory-II; gBDI = General Depression Subscale; aBDI = Anhedonia Depression Subscale; QLES-Q-SF = Quality of Life Enjoyment and Satisfaction Questionnaire-Short Form; PSC = Physical Symptom Checklist; DOCS = Dimensional Obsessive-Compulsive Scale; STAI = State-Trait Anxiety Inventory; OCTCDQ = Obsessive-Compulsive Trait Core Dimensions Questionnaire; PROMIS = Patient-Reported Outcomes Measurement Information System (T-scores).

### Analysis

Our analysis proceeded in three stages, mirroring our primary study objectives: (1) identification of data-driven clusters of general distress, (2) validation of these clusters as a distinct, multidimensional construct of general distress with convergent and discriminant validity, and (3) examination of their associations with anhedonia.

### Cluster Identification and Classification

First, k-means clustering was employed to identify data-driven clinical profiles of distress. The input features consisted of 10 key clinical variables described in the Measures section, which were standardized using *StandardScaler* prior to clustering. Multiple random initializations were performed to ensure stability of the solution. The optimal number of clusters (*k*) was determined by fit indices (evaluating the elbow method and average silhouette scores for k ranging from 2 to 10) (James et al., 2013) and clinical interpretability.

To characterize the empirically derived clusters, a series of nonparametric Kruskal-Wallis tests were conducted to compare the groups on the ten clinical variables used as input features.

For variables showing a significant overall difference, post hoc Dunn’s tests with a Bonferroni correction were performed to examine specific pairwise contrasts. Effect sizes were calculated using the rank-biserial correlation (Cureton, 1956).

### Validation of the General Distress Construct

To validate that the cluster solution of the general distress construct is not driven by a single symptom domain, we compared our k-means cluster assignments against three separate, data-driven stratifications based on unidimensional symptom severity (OCD, depression, and anxiety). Severity tiers ("High", "Intermediate", "Low") for the DOCS, gBDI, and PROMIS Anxiety scores were created based on the sample’s distribution (scores within one standard deviation of the median defined the "Intermediate" tier). Agreement between the multidimensional clusters and these unidimensional tiers was quantified using the Adjusted Rand Index (ARI) (Yin et al., 2024). Lower ARI values were interpreted as evidence of discriminant validity, indicating that the distress clusters captured variance beyond any single symptom domain.

### Relations of General Distress to Anhedonia

We conducted a series of analyses to evaluate the relationship between general distress and anhedonia. First, we used Chi-square tests and Cramér’s *V* to compare the strength of association between clinically significant anhedonia. Second, we used a hierarchical logistic regression to test whether cluster membership explained unique variance in anhedonia status, above and beyond the contribution of depression (gBDI) or OCD severity.

Finally, to shift the focus from *whether* anhedonia was present to *how* it manifests, we conducted a detailed item-level analysis of the 14 SHAPS items. For each cluster, we calculated the mean dichotomous anhedonic response rate and the mean continuous Likert score for each item. Non-parametric Kruskal-Wallis tests were used to assess for significant differences in item-level endorsement across the three clusters, providing a granular view of hedonic capacity.

All analyses were performed using Python (Van Rossum & Drake, 2009) with libraries including pandas (McKinney, 2010), scikit-learn (Pedregosa et al., 2011), and scipy.stats (Virtanen et al., 2020). Visualizations were generated using Matplotlib (Hunter, 2007) and Seaborn (Waskom, 2021).

### Sample Size Considerations

Minimum sample size requirements for cluster analysis depend on several factors, including the number and discriminability of input features, the degree of separation between clusters, and their relative sizes (Dalmaijer et al., 2022). These parameters cannot be fully determined a priori; however, simulation studies suggest that stable and replicable cluster solutions are typically achieved when subgroup sizes exceed approximately 20–30 participants, assuming moderate separation among clusters. Given our sample of 227 participants, we anticipated adequate power to detect at least 7 different groups of general distress if the clusters were relatively equal in size and moderately separated, although smaller or less distinct subgroups may not be reliably identified.

## Results

### Demographics

The final study sample comprised 227 adult participants. The mean age of participants for whom age data were available (*n* = 197) was 30.47 years (*SD* = 11.20). Information regarding sex assigned at birth was available for 209 participants; the majority identified as female (n = 145, 69.4%), 27.8% (*n* = 58) identified as male, 2.4% (*n* = 5) preferred not to disclose, and 0.5% (*n* = 1) identified as non-binary/other. Data on marital status were available for 201 participants. The largest proportion reported being single (*n* = 127, 63.2%), followed by married (*n* = 39, 19.4%), and those in other domestic partnerships (*n* = 20, 10.0%). A smaller number of participants were separated or divorced (*n* = 8, 4.0%) or reported another marital status (*n* = 7, 3.5%).

The racial composition of the sample (*n* = 209 reporting) was predominantly White/Caucasian (*n* = 162, 77.5%). Other racial identities represented were Asian (*n* = 16, 7.7%), Mixed/More than One Race (*n* = 15, 7.2%), and Black/African American (*n* = 9, 4.3%). A small number of participants were reported as Unknown (*n* = 6, 2.9%) or Native American/Other (*n* = 1, 0.5%). The large majority of participants for whom data were available identified as Non-Hispanic (*n* = 186, 89.0%), with 11.0% (*n* = 23) identifying as Hispanic.

Descriptive statistics for the primary clinical measures administered are presented in Table 1. The median score on the SHAPS for participants with complete data (*N* = 227) was 0.00 (*IQR* = 1.00), with scores ranging from 0.00 to 11.00. The SHAPS demonstrated excellent reliability (α = .90). Similarly, high internal consistency was observed for the BDI-II (α = .92), gBDI (α = .91), and DOCS Total (α = .88) and its subscales (Contamination: α = .94; Responsibility/Harm: α = .91; Unacceptable Thoughts: α = .92; Symmetry/Completeness: α = .93). Likewise, the internal consistency of the STAI State (α = .91) and Trait (α = .92) inventories, the OCTCDQ Total (α = .93) and its Harm (α = .91) and Incompleteness (α = .92) subscales were excellent. The QLES-Q-SF Total Score (α = .84), PSC Total Score (α = .83), and aBDI (α = 0.77) also demonstrated high internal consistency.

### Stage 1: Identification of Distress Profiles

Determination of the optimal number of clusters was guided by the elbow method, average silhouette scores (Figure 1), and clinical utility. We examined multiple solutions; we present detailed metrics for *k* = 2 and *k* = 3. While the *k* = 2 solution yielded a slightly higher average silhouette score than *k* = 3 (0.25 vs 0.17), indicating stronger mathematical separation, the *k* = 3 solution was ultimately chosen because it provided the best balance of statistical rigor and clinical interpretability. A full comparison demonstrating the superiority of the *k* = 3 solution is detailed in our cluster validation analysis and the Supplement.

**Figure 1:**
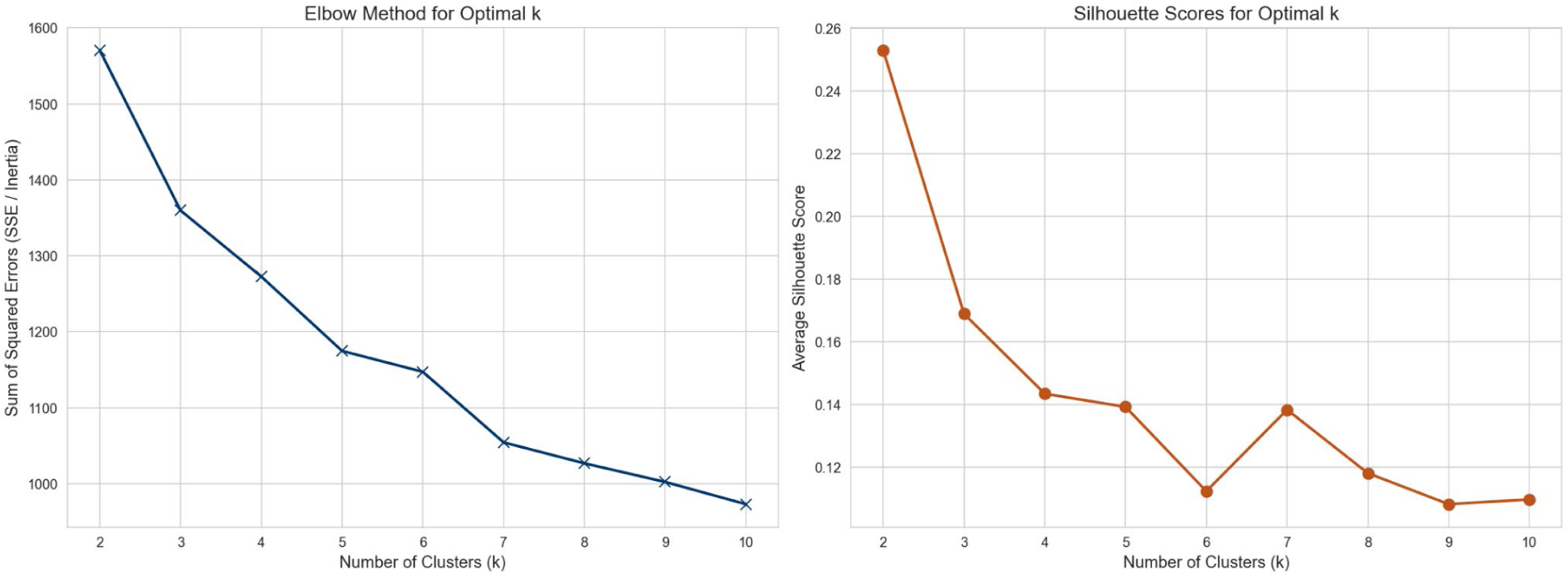
Elbow Method and Silhouette Scores for Optimal Cluster Selection

Following cluster selection, the following profiles were defined:

● **Cluster 1** ("High Global Distress and OCD"; HGD-OCD): The 62 individuals in the HGD-OCD cluster displayed a profile of pervasive distress, exhibiting the highest raw median scores on measures of depression (gBDI Mdn = 28.00, IQR = 10.25), physical symptoms (PSC Mdn = 44.00, IQR = 9.60), state anxiety (STAI State Mdn = 61.00, IQR = 6.75), trait anxiety (STAI Trait Mdn = 67.00, IQR = 6.75), and OCD symptom severity (OCTCDQ Mdn = 79.00, IQR = 9.60). Correspondingly, this group had the lowest quality of life (reversed QLES-Q-SF Mdn = 27.00, IQR = 8.00).
● **Cluster 2** ("Intermediate Overall Severity"; IOS): 100 people presented with an intermediate psychopathological profile in the IOS cluster. Raw median scores for depression (gBDI Mdn = 15.00, IQR = 8.25), OCD symptoms (OCTCDQ Mdn = 69.80, IQR = 11.60), anxiety (STAI State Mdn = 52.00, IQR = 10.00; STAI Trait Mdn = 57.00, IQR = 9.25), and physical symptoms (PSC Mdn = 36.50, IQR = 8.25) generally fell between those of the HGD-OCD and LSP clusters. Similarly, quality of life for this group (reversed QLES-Q-SF Mdn = 16.00, IQR = 9.00) was positioned between the other two extremes.
● **Cluster 3** ("Low Symptom Profile"; LSP): The 65 participants in the LSP cluster consistently showed the lowest raw median scores across all measures of psychopathology despite receiving a confirmed diagnosis of OCD (e.g., gBDI Mdn = 7.00, IQR = 6.00; OCTCDQ Mdn = 58.00, IQR = 18.00; STAI State Mdn = 41.00, IQR = 13.00; STAI Trait Mdn = 45.00, IQR = 10.00) and the highest quality of life (reversed QLES-Q-SF Mdn = 8.00, IQR = 8.00).

Standardized mean profiles of these three clusters on each input variable are depicted in Figure 2, and pairwise statistical comparisons of the input variables are presented in Table 3. While the HGD-OCD and LSP clusters showed consistently high and low scores respectively, it is notable that this pattern was not a simple linear gradation across all variables. For instance, scores on the DOCS Symmetry subscale did not significantly differ between adjacent clusters, suggesting that symmetry-related symptoms may be less tightly coupled with the overall distress construct.

**Figure 2:**
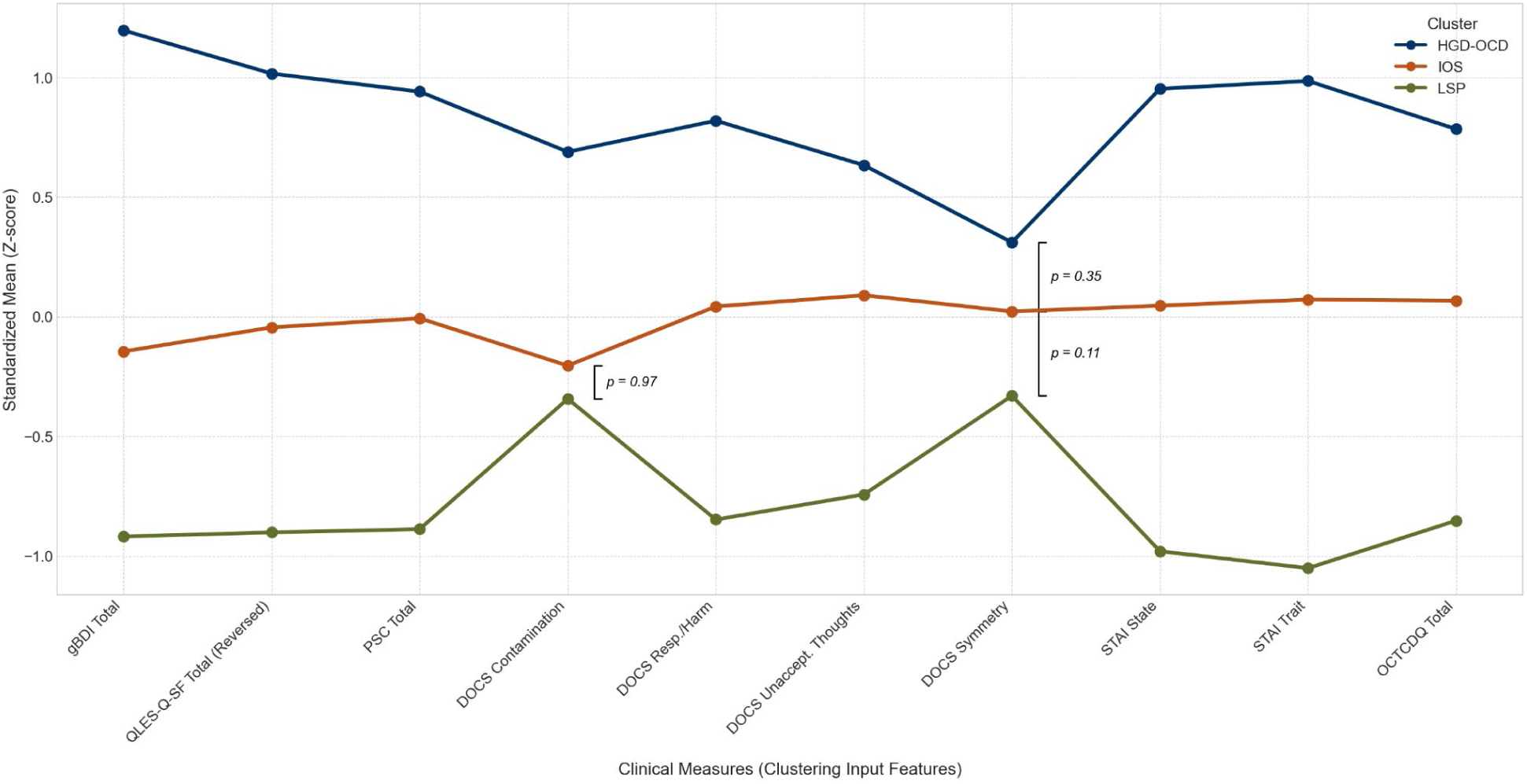
Standardized Mean Profiles for k = 3 Clusters. *Note:* Standardized (z-score) means are displayed to allow for visual comparison across measures with different scales. gBDI = General Depression Subscale; QLES-Q-SF = Quality of Life Enjoyment and Satisfaction Questionnaire-Short Form; PSC = Physical Symptom Checklist; DOCS = Dimensional Obsessive-Compulsive Scale; STAI = State-Trait Anxiety Inventory; OCTCDQ = Obsessive-Compulsive Trait Core Dimensions Questionnaire. All measures differ significantly among the three clusters (Kruskal-Wallis, *p* < .005). Brackets indicate non-significant pairwise comparisons (*p* > .05) between adjacent clusters after Bonferroni correction; all other pairwise comparisons were statistically significant. See Table 3 for more details.

**Table 3:**
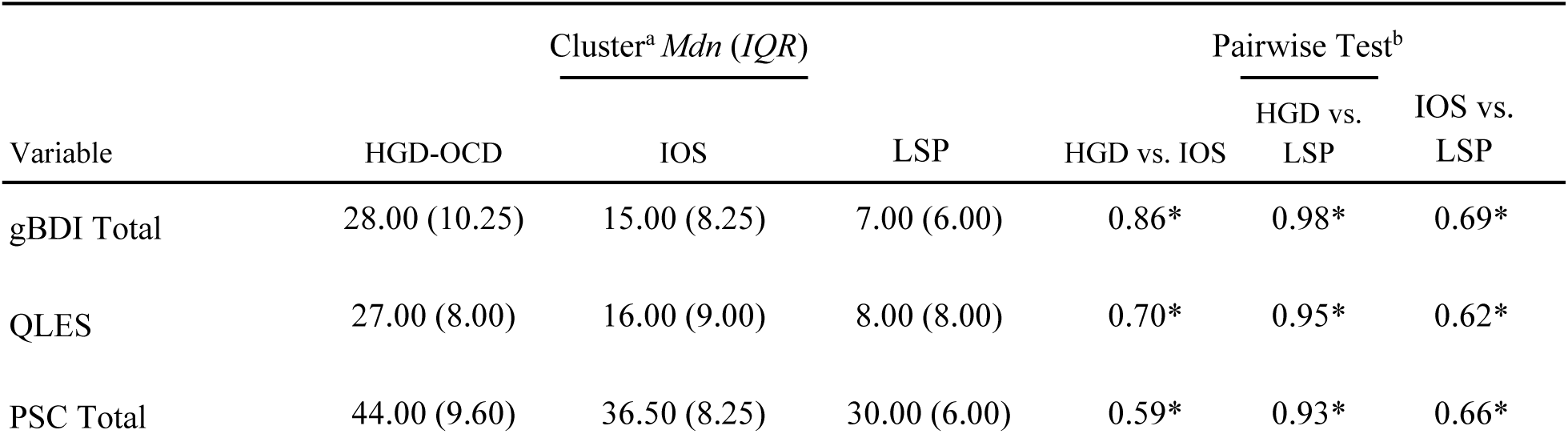
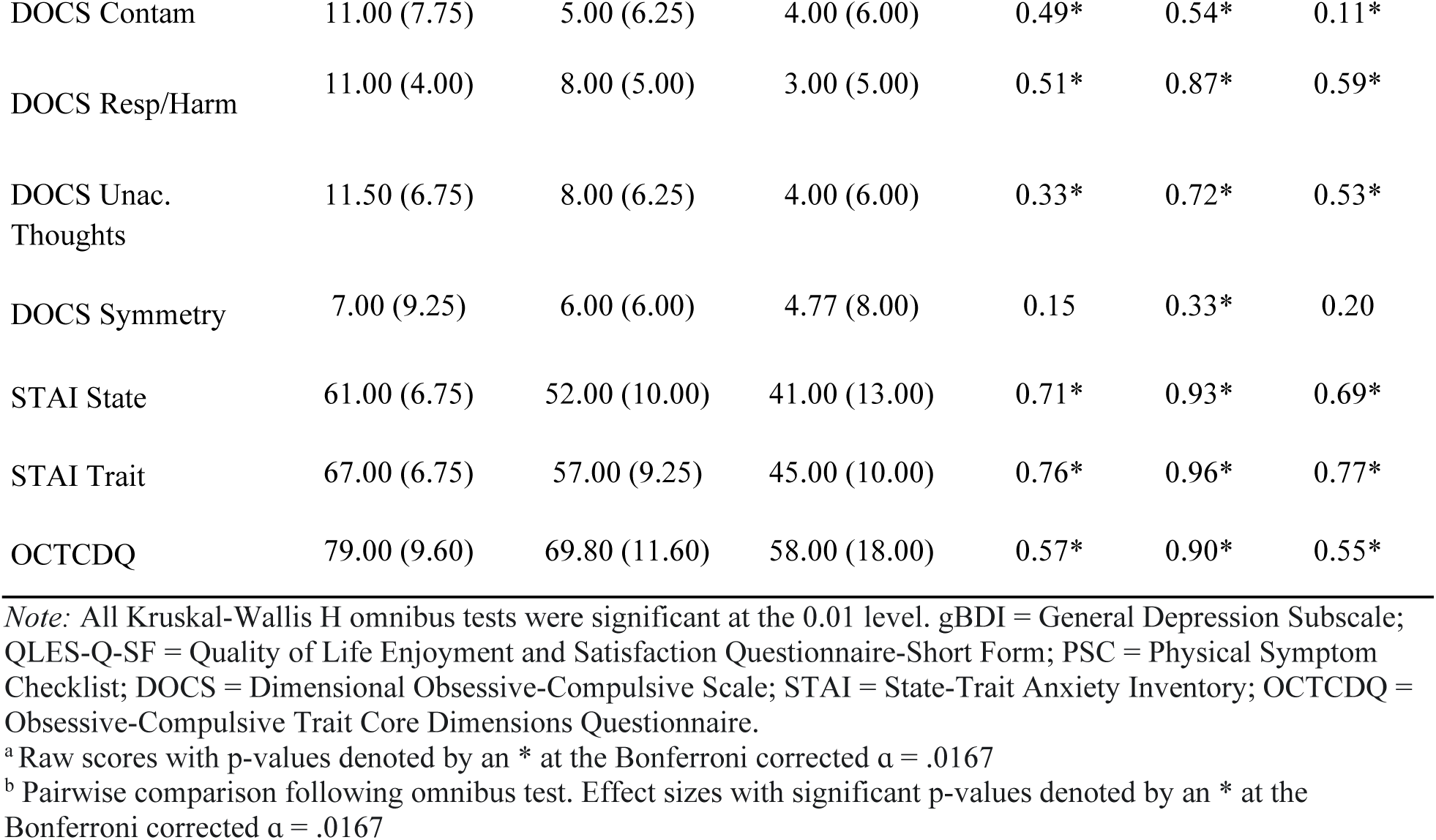
Statistical Comparison of Clustering Variables.

| Variable | Cluster<sup>a</sup> *Mdn (IQR)* |  |  | Pairwise Test<sup>b</sup> |  |  |
| --- | --- | --- | --- | --- | --- | --- |
|  | HGD-OCD | IOS | LSP | HGD vs. IOS | HGD vs. LSP | IOS vs. LSP |
| gBDI Total | 28.00 (10.25) | 15.00 (8.25) | 7.00 (6.00) | 0.86* | 0.98* | 0.69* |
| QLES | 27.00 (8.00) | 16.00 (9.00) | 8.00 (8.00) | 0.70* | 0.95* | 0.62* |
| PSC Total | 44.00 (9.60) | 36.50 (8.25) | 30.00 (6.00) | 0.59* | 0.93* | 0.66* |
| DOCS Contam | 11.00 (7.75) | 5.00 (6.25) | 4.00 (6.00) | 0.49* | 0.54* | 0.11* |
| DOCS Resp/Harm | 11.00 (4.00) | 8.00 (5.00) | 3.00 (5.00) | 0.51* | 0.87* | 0.59* |
| DOCS Unac.<br>Thoughts | 11.50 (6.75) | 8.00 (6.25) | 4.00 (6.00) | 0.33* | 0.72* | 0.53* |
| DOCS Symmetry | 7.00 (9.25) | 6.00 (6.00) | 4.77 (8.00) | 0.15 | 0.33* | 0.20 |
| STAI State | 61.00 (6.75) | 52.00 (10.00) | 41.00 (13.00) | 0.71* | 0.93* | 0.69* |
| STAI Trait | 67.00 (6.75) | 57.00 (9.25) | 45.00 (10.00) | 0.76* | 0.96* | 0.77* |
| OCTCDQ | 79.00 (9.60) | 69.80 (11.60) | 58.00 (18.00) | 0.57* | 0.90* | 0.55* |

*Note:* All Kruskal-Wallis H omnibus tests were significant at the 0.01 level. gBDI = General Depression Subscale; QLES-Q-SF = Quality of Life Enjoyment and Satisfaction Questionnaire-Short Form; PSC = Physical Symptom Checklist; DOCS = Dimensional Obsessive-Compulsive Scale; STAI = State-Trait Anxiety Inventory; OCTCDQ = Obsessive-Compulsive Trait Core Dimensions Questionnaire.
<sup>a</sup> Raw scores with p-values denoted by an \* at the Bonferroni corrected $\alpha = .0167$
<sup>b</sup> Pairwise comparison following omnibus test. Effect sizes with significant p-values denoted by an \* at the Bonferroni corrected $\alpha = .0167$

### Stage 2: Validation of the Clusters as a Multidimensional Construct

A central hypothesis of this study was that the empirically derived clusters represent a separate multidimensional construct of general distress, not simply a proxy for unidimensional symptom severity. To test this, we compared our cluster assignments against the data-driven severity tiers for OCD (DOCS), depression (gBDI), and anxiety (PROMIS Anxiety) presented in Table 4.

**Table 4:**
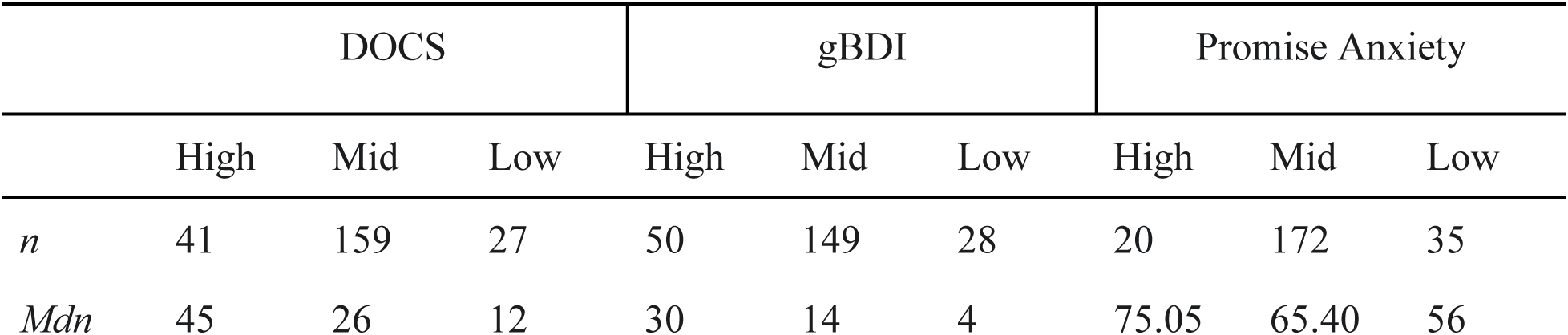
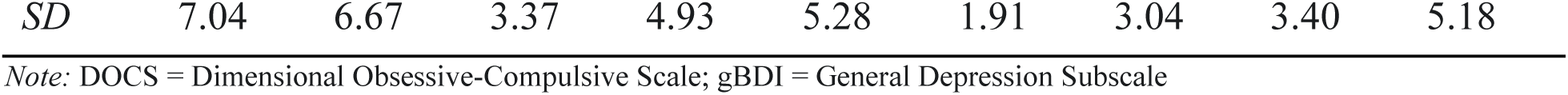
Characteristics of Data-Driven Unidimensionality Severity Tiers.

The agreement between our multidimensional clusters and these unidimensional tiers was quantified using the Adjusted Rand Index (ARI). The analysis revealed a pattern of agreement based on point estimates. The overlap was numerically highest with depression tiers (ARI = 0.28), followed by anxiety (ARI = 0.17), and finally a near-random relationship with OCD severity tiers (ARI = 0.11). This pattern suggests that the identified clusters capture broader variance. However, a formal comparison using a non-parametric bootstrap analysis indicated that these differences were not statistically significant (e.g., 95% CI for the difference between depression and anxiety ARI: [-0.0199, 0.2121]), suggesting that while a trend exists, the cluster solution’s alignment with depression is not definitively stronger than its alignment with anxiety or OCD in this sample. The heatmaps in Figure 3 depict these findings.

**Figure 3:**
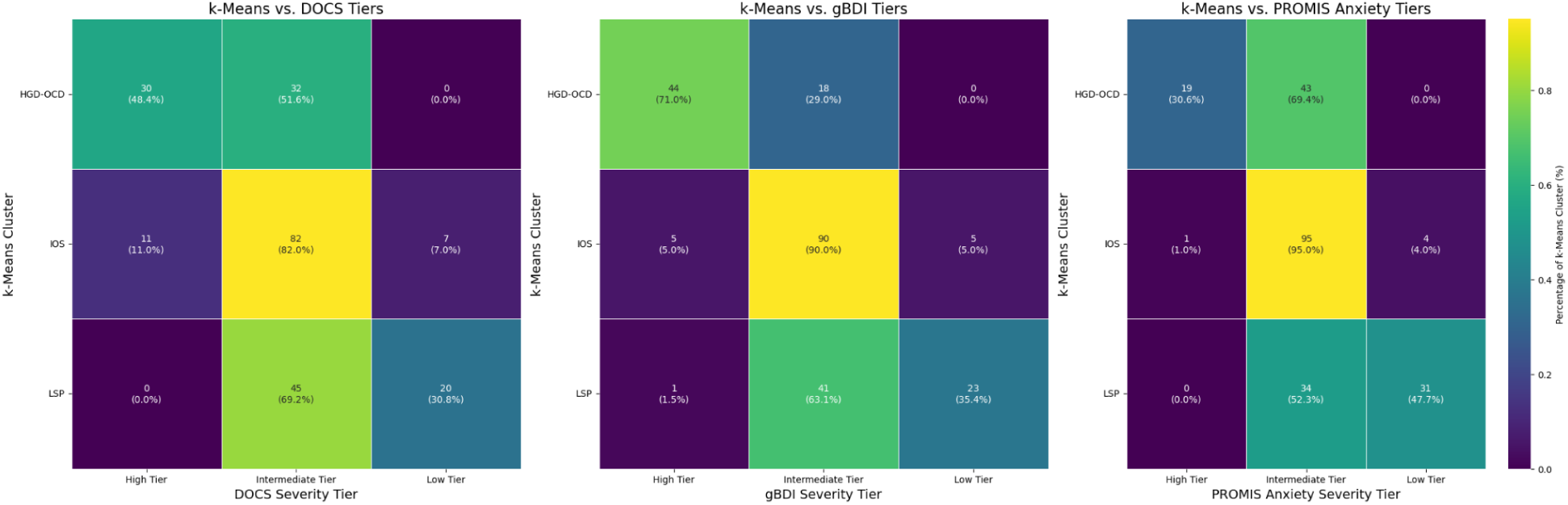
Overlap between the k-Means Clusters and Data Driven-Derived Severity Tiers from the Dimensional Obsessive Compulsive Scale, the general Beck Depression Inventory II, and the PROMIS Anxiety. Color intensity is based on the percentage contained by each k-means cluster).

While the clusters showed a qualitatively stronger alignment with depression severity—for example, 71.0% of the HGD-OCD cluster fell into the high gBDI tier—the discordance highlights their unique, multidimensional nature. Over half of the HGD-OCD cluster (51.6%) did not fall into the high OCD severity tier, demonstrating that a patient’s overall distress level is not synonymous with their primary diagnosis severity. In contrast, the IOS cluster remained highly consistent across measures, with 82.0% to 95.0% of its members occupying the intermediate tier for both OCD and depression, solidifying its identity as a robust moderate-distress group. Together, these results validate the clusters as a novel construct of general distress.

### Stage 3: Distress Profiles Predict Anhedonia Presence, Severity, and Nature

Having identified and validated the distress profiles, we proceeded to test our primary hypotheses regarding their relationship with anhedonia—a construct intentionally excluded from the clustering process.

### Anhedonia Severity Differs Significantly Across Clusters

As shown in Figure 4 and Table 5, the three clusters differed significantly on all external constructs. On the primary anhedonia measure (SHAPS), there was a gradation in severity *H*(2) = 18.75, *p* < .001: the HGD-OCD cluster reported the highest levels, followed by the IOS cluster, then the LSP cluster, where anhedonia was virtually absent. A similar monotonic pattern was observed for the anhedonia subscale of the BDI-II (aBDI) and its individual items (Table 5).

**Figure 4:**
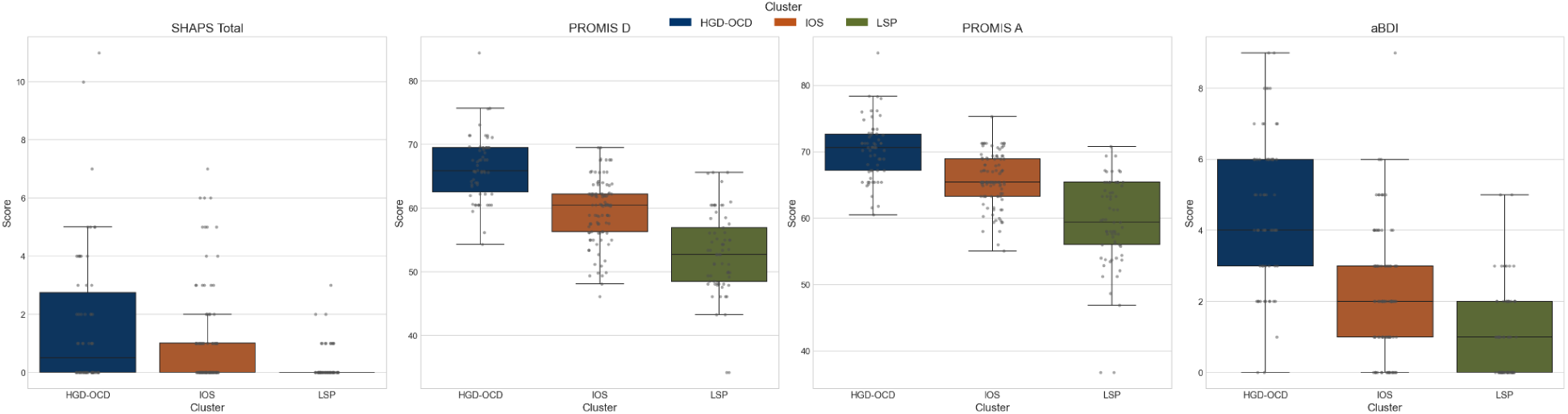
k = 3 Raw Score Cluster Comparison on External Clinical Variables. HGD OCD (High Global Distress OCD cluster), IOS (Intermediate Overall Severity cluster), LSP (Low Symptom Profile cluster).

**Table 5:** Comparison of *k* = 3 Clusters on External Clinical Variables.

| Feature | HGD<br>OCD<sup>a</sup> | IOS<sup>b</sup> | LSP<sup>c</sup> | Stat<sup>f</sup> | $p$ |
| --- | --- | --- | --- | --- | --- |
| PROMIS D<br>*Mdn (IQR)* | 65.85<br>(7.00) | 60.50<br>(5.90) | 52.70<br>(8.40) | 109.3 | <.001 |
| PROMIS A<br>*Mdn (IQR)* | 70.60<br>(5.45) | 65.40<br>(5.60) | 59.40<br>(9.40) | 92.13 | <.001 |
| aBDI Total<br>*Mdn (IQR)* | 4.00<br>(3.00) | 2.00<br>(2.00) | 1.00<br>(2.00) | 80.96 | <.001 |
| aBDI 4<br>% $\geq 2$ | 53.2%<br>(33/62) | 13.0%<br>(13/100) | 1.5%<br>(1/65) | 58.1 | <.001 |
| aBDI 12<br>% $\geq 2$ | 48.4%<br>(30/62) | 10.0%<br>(10/100) | 1.5%<br>(1/65) | 54.91 | <.001 |
| aBDI 21<br>% $\geq 2$ | 50.0%<br>(31/62) | 21.0%<br>(21/100) | 3.1%<br>(2/65) | 39.3 | <.001 |

*Note:* HGD OCD (High Global Distress OCD cluster), IOS (Intermediate Overall Severity cluster), LSP (Low Symptom Profile cluster)

### Cluster Membership is a Superior Predictor of Anhedonia

We next compared the strength of association between clinically significant anhedonia and our multidimensional clusters versus the unidimensional severity tiers. As shown in Table 6, the point estimate for the association for our multidimensional clusters (Cramér’s *V* = 0.26, *p* < .001) was similar to that of depression severity (*V* = 0.26, *p* < .001). These values were numerically higher than the association with anxiety (*V* = 0.19, *p* = .021) or OCD severity (*V* = 0.14, *p* = .113). Follow-up bootstrap analyses to test these differences revealed they were not statistically significant (95% CI for clusters vs anxiety: [-0.03, 0.21]; 95% CI for clusters vs OCD: [< 0.01, 0.22]). This indicates that the explanatory power of our multidimensional clusters is on par with, but not statistically superior to, the unidimensional tiers in explaining anhedonia.

**Table 6:** Association with Clinically Significant Anhedonia and Unidimensional Severity Tiers.

| Measure | $N$ | $\chi^2$ | df | $p$ | Cramér's $V$ |
| --- | --- | --- | --- | --- | --- |
| Cluster | 33/137 | 15.35 | 2 | $< .001$ | 0.26 |
| gBDI | 15 / 33 | 15.11 | 2 | $< .001$ | 0.26 |
| PROMIS<br>Anxiety | 6 / 33 | 7.74 | 2 | .021 | 0.19 |
| DOCS | 9 / 33 | 4.37 | 2 | .113 | 0.14 |

Note: gBDI = General Depression Subscale; DOCS = Dimensional Obsessive-Compulsive Scale

To test whether the distress profiles explain anhedonia beyond its primary clinical correlates, we performed a hierarchical logistic regression. Adding OCD severity (DOCS total score) to a baseline model containing only depression severity (gBDI) did not significantly improve model fit χ²(1) = 0.13, *p* = .719. In the final step, we added the cluster assignments to the standard clinical model. This resulted in a statistically significant improvement in model fit χ²(2) = 7.95, *p* = .019. Critically, in this full model, cluster membership emerged as the sole significant predictor. Compared to the LSP group, participants in the HGD-OCD cluster had significantly higher odds of clinically significant anhedonia (OR = 17.24, *p* = .037), as did participants in the IOS cluster (OR = 11.49, *p* = .027). Notably, with cluster membership accounted for, the effects of both gBDI (*p* = .369) and DOCS (*p* = .484) were rendered non-significant. Full logistic regression results are presented in Supplement 2.

### Anhedonia Manifests in Distinct, Facet-Specific Profiles

Finally, to understand how anhedonia manifests across clusters, we examined the relationship between item-level responses on the SHAPS and cluster membership (Figure 5).

**Figure 5:**
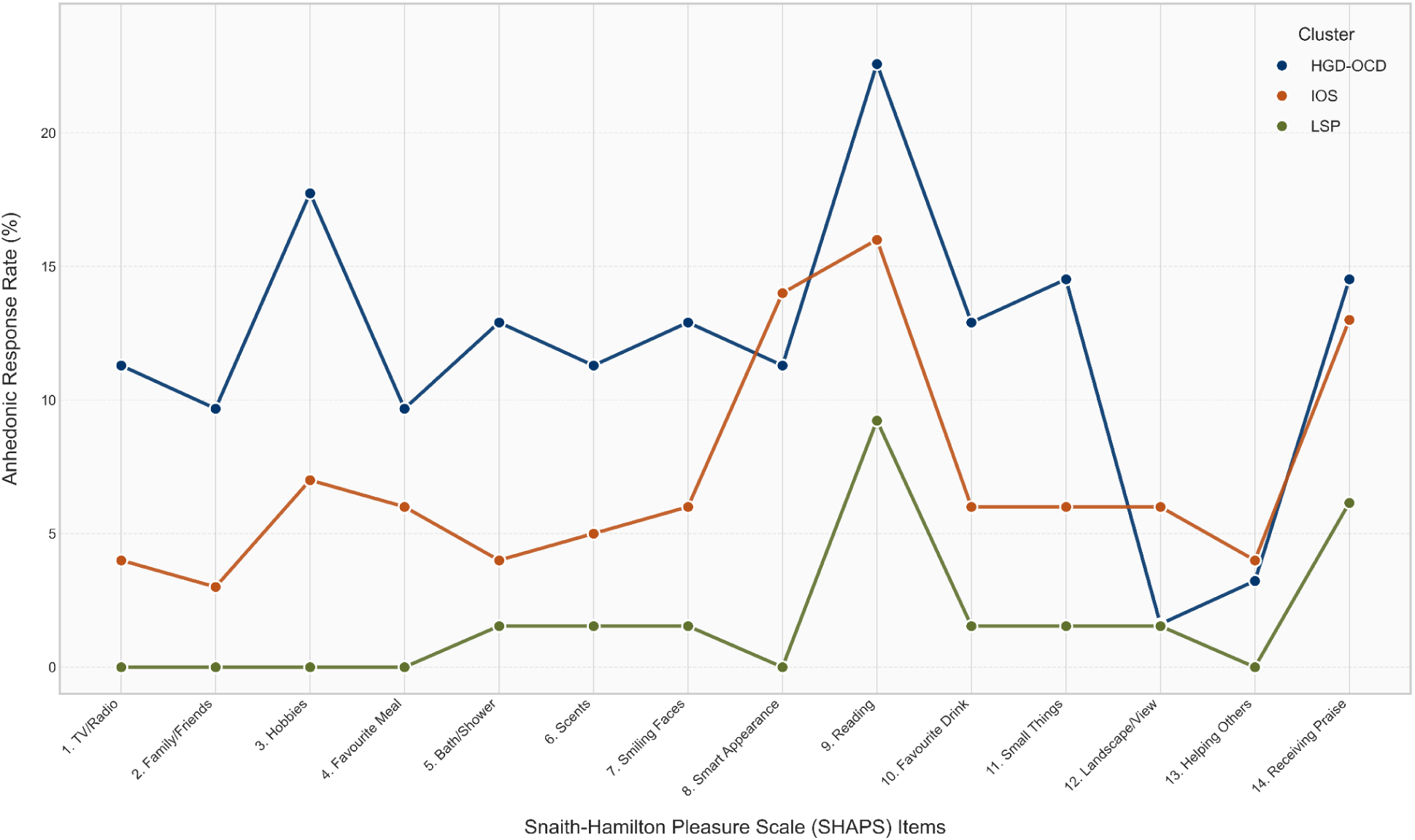
Item-Level Anhedonia Profiles Across OCD Clusters

The HGD-OCD cluster exhibited the most broadly impaired hedonic capacity, with the highest anhedonic response rates across nearly all items, particularly for activities like "Reading" (22.6%) and "Hobbies" (17.7%). The IOS cluster displayed a complex, varied pattern. Its profile resembled the high-distress group on items related to social contribution ("Helping Others"), but was closer to the low-distress group on items like enjoying time with "Family/Friends." This illustrates that the IOS profile is not merely a midpoint, but a distinct clinical entity with specific domains of both preserved and impaired hedonic capacity. The LSP cluster showed a consistently preserved hedonic capacity, with near-zero anhedonic response rates across most items.

## Discussion

This study used a cluster analysis to identify empirically derived distress profiles in individuals with OCD. The analysis revealed a High Global Distress and OCD (HGD-OCD) group, an Intermediate Overall Severity (IOS) group, and a Low Symptom Profile (LSP) group. A central finding is the modest agreement between these clusters and unidimensional severity tiers for depression (ARI = 0.28), anxiety (ARI = 0.17), and OCD (ARI = 0.11), which supports our hypothesis that these profiles represent a higher-order construct of distress not reducible to any single symptom domain. This reinforces the view that univariate approaches may mask the clinical reality that individuals with similar levels of OCD and depression experience fundamentally different patterns of overall distress (Lochner & Stein, 2003, 2006).

The relationship with specific OCD symptom domains was similarly complex. While most measures scaled with overall distress, DOCS Symmetry/Completeness scores did not. This non-linear pattern, which would be obscured by a simple severity model, suggests that symmetry-related symptoms may have a more stable presentation across distress levels, hinting at distinct underlying mechanisms (e.g., a "just right" feeling rather than reward deficit) that warrant further investigation (Schwartz, 2018; Stern et al., 2022).

The hierarchical logistic regression formally confirmed the unique explanatory power of the distress profiles. The analysis revealed that when depression severity was used as a baseline predictor of anhedonia, adding OCD severity offered no additional information. However, the final model including our cluster profiles not only provided a significantly better fit but also rendered the individual effects of both depression and OCD severity non-significant. This is a key finding: It suggests that the holistic pattern of a patient’s distress—as captured by the HGD-OCD and IOS profiles—is a more robust and fundamental predictor of anhedonia than the severity of any single comorbid symptom. This supports the conceptualization of anhedonia in OCD not merely as a byproduct of depression, but as an emergent property of the broader, more complex factor of general distress.

For instance, agreement between distress and depression tiers was modest (ARI = 0.28): 29% of participants in the HGD-OCD group did not meet high-tier depression criteria, whereas approximately 63% of those in the LSP group reported intermediate levels of depression.

Agreement with OCD-based tiers was even lower (ARI = 0.11), with 51% of the high-distress group and 61% of the low-distress group reporting intermediate OCD symptoms. Although the prevalence of clinically significant anhedonia increased monotonically across the distress profiles—from the LSP to IOS to HGD-OCD groups—the item-level patterns revealed a more nuanced picture. The SHAPS analyses identified three distinct configurations of hedonic capacity that mirrored the cluster-level findings. The HGD-OCD profile was characterized by a broad impairment in hedonic capacity, whereas the LSP profile was defined by its preservation. Most critically, the IOS profile was not merely an intermediate between high and low distress groups. This profile, identifiable only through clustering, serves as the clearest illustration of the limitations of unidimensional severity measures. For instance, while a simple depression score might classify these 100 individuals as ’moderate,’ it would obscure the clinical detail that their hedonic capacity encompasses both profound impairment (e.g., in social contribution) and preserved function (e.g., enjoying family/friends). This differentiation provides the strongest support for a multidimensional approach, demonstrating that anhedonia is not a monolithic construct but comprises specific facets that are differentially expressed depending on a patient’s overarching clinical profile. This pattern also suggests potential mechanistic distinctions, such as motivational versus consummatory deficits, that require further investigation (Shankman et al., 2014). For instance, within the IOS group, elevated anhedonia on items related to social contribution ("Helping Others") resembled the high-distress group, suggesting social anhedonia may be sensitive to increases in distress (King & Zaboski, 2024; Moore, 2021; Xia et al., 2019). In contrast, responses on the Family/Friends item were closer to those of the low-distress group, indicating that hedonic impairment in this intermediate tier is domain-specific rather than global.

These findings suggest that certain aspects of hedonic experience may begin to decline earlier than others as overall distress increases. Specifically, enjoyment derived from effortful or socially mediated rewards appears more vulnerable to rising distress, whereas pleasure from more automatic or sensory activities (e.g., scents, bathing/showering) remains relatively preserved until distress becomes severe. This graded erosion of hedonic capacity supports the view that global distress in OCD progressively constrains hedonic capacity, particularly for complex or affiliative forms of pleasure.

## Clinical Implications

The identification of these distinct profiles allows clinicians to begin moving beyond a one-size-fits-all diagnostic label ("OCD") toward a more nuanced, data-driven approach to understanding and potentially treating patient subgroups (Fernandes et al., 2017; Zaboski & Bednarek, 2025). First, clinicians should not assume a uniform presence or absence of anhedonia in OCD and should consider routine, facet-specific assessment. For individuals presenting with the HGD-OCD profile, the combination of high anhedonia and pervasive distress suggests that standard ERP may need to be augmented with interventions targeting broader affective symptoms, such as behavioral activation (Ciharova et al., 2021; Cuijpers et al., 2007). For those fitting the IOS profile, a targeted approach focused on specific domains of reduced pleasure (e.g., social engagement, hobbies) may be most effective. Second, the clustering analysis could help identify patients from the outset who may need modified treatment protocols. Individuals in the HGD-OCD cluster, for example, may be more likely to benefit from treatments that address potential challenges in motivation and engagement, as suggested by recent work showing that patients with high anhedonia may require more sessions of exposure therapy to achieve similar gains (Mattoni et al., 2025).

## Limitations and Future Directions

This study has several limitations. First, its cross-sectional design precludes inferences about causality or the developmental trajectory of these distress profiles and their relationship with anhedonia. Longitudinal studies are needed to track how these profiles evolve and whether baseline anhedonia levels within these profiles predict treatment response and long-term outcomes. While the SHAPS and BDI-II items provided valuable initial insights into global and some facet-specific anhedonia, differentiating between anticipatory anhedonia (deficits in the motivation or expectation of pleasure) and consummatory anhedonia (deficits in the experience of pleasure during an activity) would allow for a more precise characterization of pleasure deficits within the identified clusters (Chapman et al., 1976; Gard et al., 2007; Li et al., 2019; Moore, 2021). Third, the sample was recruited from a single research clinic, which may limit generalizability. Replication in more diverse clinical and community samples is crucial. Fourth, the cluster analysis itself, while data-driven, involves decisions (e.g., number of clusters) that can influence outcomes. Future studies could explore the stability of these clusters using different methods (e.g., hierarchical clustering, Gaussian Mixture Models, Latent Profile Analysis, in different samples, and consider variables that might mitigate distress (e.g., coping/resilience).

Lastly, while our sample size was sufficient to identify three reasonably well-sized and distinct clusters, future research with larger samples could identify smaller subgroups.

## Conclusion

This study demonstrates the utility of a multidimensional approach for understanding the interplay between general distress and anhedonia in individuals with OCD. By first identifying and validating holistic distress profiles, our approach moved beyond simple gradations of severity to uncover clinically meaningful complexity. The identification of distinct subgroups with varying degrees and, critically, different patterns of anhedonia underscores the importance of looking beyond a single diagnosis/comorbidity, or using univariate measures within linear analyses. Further investigation into these profiles may pave the way for more targeted and effective interventions, tailored not just to a patient’s primary diagnosis, but to their complete clinical presentation.

## Data Availability

All data produced in the present study are available upon reasonable request to the authors.

## Supplement 1

A supplemental analysis was run to quantitatively explore the choice of *k* = 2 vs *k* = 3 clusters. The *k* = 2 solution delineates a global ’High Distress’ (HGD-OCD) versus ’Low Distress’ profile (See Fig S1), where the profiles are largely parallel and do not capture the nuanced interactions between symptom domains seen in the *k* = 3 solution. While mathematically justified by the highest silhouette score, the *k* = 2 solution obscures the clinically relevant ‘Intermediate Overall Severity’ (IOS) group identified in the main analysis.

**Figure S1:**
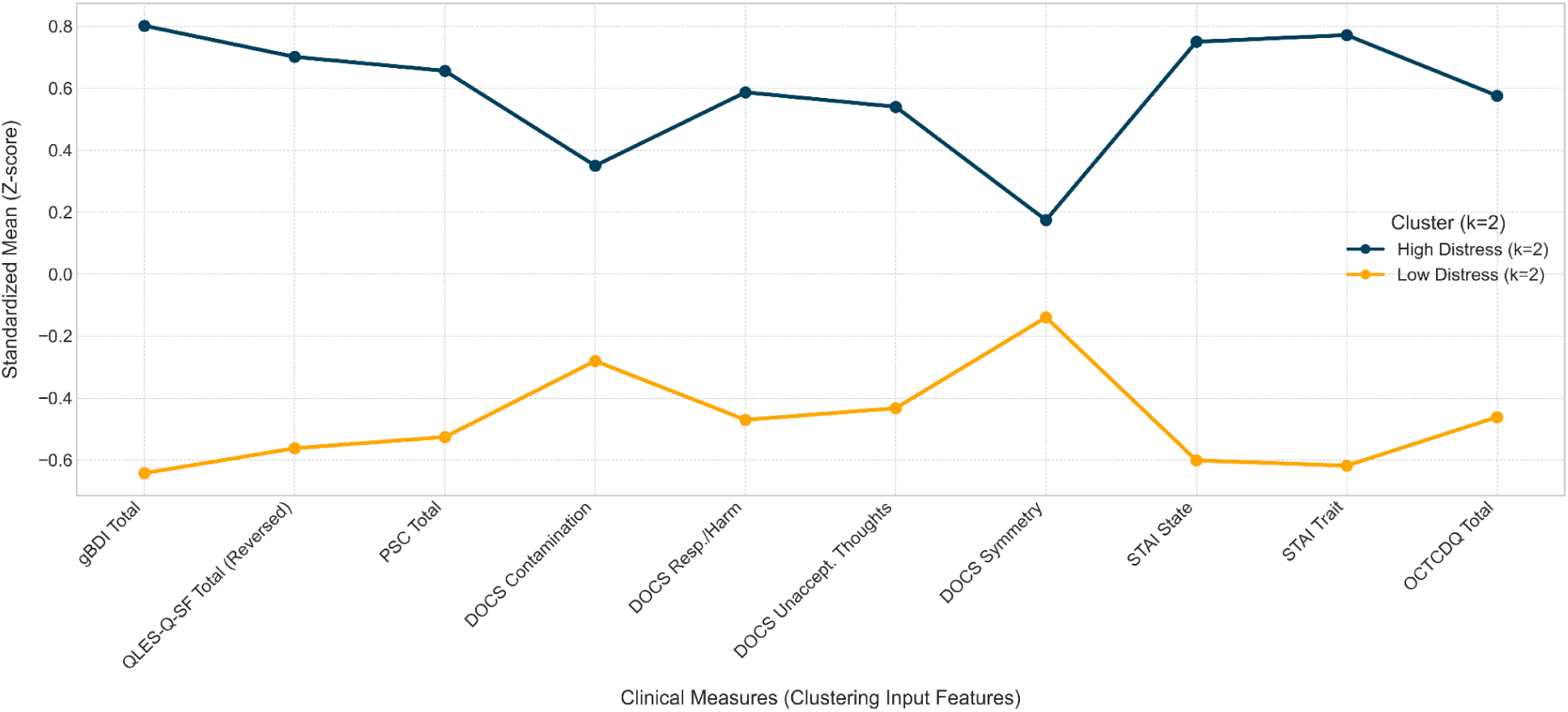
Standardized Mean Profiles for k = 2 Clusters. *Note:* Standardized (z-score) means are displayed to allow for visual comparison across measures with different scales. gBDI = General Depression Subscale; QLES-Q-SF = Quality of Life Enjoyment and Satisfaction Questionnaire-Short Form; PSC = Physical Symptom Checklist; DOCS = Dimensional Obsessive-Compulsive Scale; STAI = State-Trait Anxiety Inventory; OCTCDQ = Obsessive-Compulsive Trait Core Dimensions Questionnaire.

Crossstabulation data demonstrated that the *k* = 2 solution achieved its simplicity by partitioning IOS group members into the "High" and "Low" distress profiles. Sixty one percent of the intermediate group was classified as "Low Distress." However, this represents a significant loss of clinical information as these individuals are more distressed than the LSP group, but with this solution, are now indistinguishable from them. Likewise, thirty-nine percent of the members in the intermediate group were absorbed into the "High Distress" cluster. This also obscures their profile, lumping them in with the most severely ill patients, even though their profile is less severe.

**Table S1:** Crosstabulation of *k* = 3 and *k* = 2 Cluster Assignment.

| $k = 3$ Cluster | Assigned to<br>'High Distress' | Assigned to 'Low<br>Distress' | Total |
| --- | --- | --- | --- |
| **HGD-OCD** | 62 (100.00%) | 0 (0.00%) | 62 |
| **IOS** | 39 (39.00%) | 61 (61.00%) | 100 |
| **Low** | 0 (0.00%) | 65 (100.00%) | 65 |

*Note:* This table displays the number of individuals from each of the three final clusters ( $k = 3$ solution) and shows how they were assigned into the two clusters of the simpler $k = 2$ solution. Values represent the raw count of participants, with the row-wise percentage in parentheses.

Overall, our sensitivity analysis confirmed that the k = 2 solution, while statistically parsimonious, achieved simplicity by partitioning 100 members of the intermediate cluster into the high and low distress profiles. This masks a clinically significant cohort and misrepresents their distinct, intermediate level of psychopathology, underscoring the necessity of the k = 3 solution for accurately capturing the heterogeneity in the sample.

## Supplement 2

This supplement presents the logistic regression results for three models: A baseline model (anhedonia predicted by gBDI), a Standard Clinical Model (anhedonia predicted by gBDI and OCD severity), and a full model (anhedonia predicted by gBDI, OCD severity, and cluster assignment). Results indicated that the k-means clusters are independent predictors of anhedonia above and beyond depression and OCD severity.

**Table S3:** Full Model Results.

| Predictor | Baseline | Standard Clinical Model | Full Model with Cluster Membership |
| --- | --- | --- | --- |
|  | *B (SE)*<br>OR [95% CI] | *B (SE)*<br>OR [95% CI] | *B (SE)*<br>OR [95% CI] |
| **Intercept** | -3.04 (0.45)***<br>0.05 [0.02, 0.12] | -3.16 (0.57)***<br>0.04 [0.01, 0.13] | -4.13 (1.12)***<br>0.02 [0.00, 0.14] |
| **gBDI** | 0.07 (0.02)***<br>1.07 [1.03, 1.11] | 0.06 (0.02)**<br>1.06 [1.02, 1.11] | 0.03 (0.03)<br>1.03 [0.97, 1.10] |
| **DOCS Total (OCD)** | - | 0.01 (0.02)<br>1.01 [0.97, 1.04] | -0.02 (0.02)<br>0.98 [0.94, 1.03] |
| **Cluster: HGD-OCD** | - | - | 2.85 (1.36)*<br>17.29 [1.19, 250.3] |
| **Cluster: IOS** | - | - | 2.44 (1.10)*<br>11.49 [1.33, 99.58] |

*Note:* Model 1 (depression only), Model 2 (Depression + OCD), Model 3 (Depression + OCD + Cluster
Assignment). *B* = log-odds coefficient; OR = odds ratio. The reference category for the cluster variable is 'LSP' (Low Symptom Profile).
\* $p < .05$ , \*\* $p < .01$ , \*\*\* $p < .001$

Likelihood ratio tests were conducted to formally compare the nested logistic regression models. First, we compared a model with both depression and OCD severity (Model 2) against a model with depression alone (Model 1). The test was not statistically significant χ²(1) = 0.13, *p* = .719, indicating that OCD severity did not improve the model. Next, we compared the full model containing cluster assignments (Model 3) against the standard clinical model with depression and OCD severity (Model 2). This comparison was statistically significant χ²(2) = 7.95, *p* = .019, demonstrating that the inclusion of the distress profiles provided an improvement in model fit. This supports the hypothesis that the clusters explain unique variance in anhedonia, even after controlling for both depression and OCD severity.

